# Applications and barriers to use of an mHealth iPhone application for self-management of chronic recurrent medical conditions: A Pilot Study

**DOI:** 10.1101/2020.04.28.20082297

**Authors:** Archana Mande, Susan L. Moore, Farnoush Banaei-Kashani, Alexander M. Kaizer, Benjamin Echalier, Sheana Bull, Michael A. Rosenberg

**Affiliations:** Clinical Research Support Team (CReST), University of Colorado Anschutz Medical Campus, Aurora, CO, USA; mHealth Impact Laboratory, University of Colorado Anschutz Medical Campus, Aurora, CO, USA; Colorado School of Public Health, University of Colorado, Aurora, CO, USA; College of Engineering and Applied Science, University of Colorado-Denver, Denver, Colorado, USA; Department of Biostatistics and Informatics, Colorado School of Public Health, University of Colorado, Aurora, CO, USA; Colorado Center for Personalized Medicine, University of Colorado Anschutz Medical Campus, Aurora, CO, USA

## Abstract

Management of chronic recurrent medical conditions (CRMC), such as migraine headaches, chronic pain and anxiety/depression, is a major challenge for modern providers. The fact that often the most effective treatments and/or preventative measures for CRMCs vary from patient to patient lends itself to a platform for self-management by patients. However, to develop such an mHealth app requires an understanding of the various applications, and barriers, to real-world use. In this pilot study with internet-based recruitment, we conducted an assessment of user satisfaction of the iMTracker iOS (iPhone) application for CRMC self-management through a self-administered survey of subjects with CRMCs. From May 15, 2019 until March 27, 2020, we recruited 135 subjects to pilot test the iMTracker application for user-selected CRMCs. The most common age group was 31–45 (48.2%), followed by under 30 (22.2%) and 46–55 (20%). There were no subjects over 75 years old completing the survey. 38.8% of subjects were college graduates, followed by 29.6% with a Master’s degree, and 25.9% with some college. No subjects had not graduated from high school, and only 2 (1.5%) did not attend college after high school. 80.7% of subjects were self-identified as Caucasian, and 90.4% as not Hispanic or Latino. The most common CRMC was pain (other than headaches) in 40% of subjects, followed by mental health in 17.8% and headaches in 15.6%. 39.3% of subjects experienced the condition multiple times in a day, 40.0% experienced the condition daily, and 14.8% experienced the condition weekly, resulting in a total of 94.1% of subjects experiencing the condition at least weekly. Among the concerns about a self-management app, time demands (54.8%) and ineffectiveness (43.7%) were the most prominent, with privacy (24.4%) and data security (25.2%) also noted. In summary, we found internet-based recruitment identified primarily Caucasian population of relatively young patients with CRMCs of relatively high recurrence rate. Future work is needed to examine the use of this application in older, underrepresented minorities, and lower socioeconomic status populations.

## Background

Chronic recurrent medical conditions (CRMCs) encompass a major proportion of the modern healthcare burden, accounting for significant costs in the form of both management and lost productive time^1^. For example, chronic migraine headaches affect ~2% of the global population^2^, and in the United States alone, cost more than $20 billion annually^1^ to manage. Chronic low back pain accounts for over 5 hours per week in lost productivity by workers, resulting in over $10 billion in lost revenue per year^1^. Mental health disorders, including depression and anxiety, accounted for 183.9 million disability-adjusted life years and 175.3 million years lived with disability worldwide^3^, with an increase of 37.6% over the years from 1990 to 2010^3^.

On a more granular level, CRMCs create a major challenge for today’s busy clinician. Although widely variable across providers and practices, the time available for a face-to-face encounter with patients continues to trend downward, despite an increase in the number of clinical items needing to be addressed^4^. As a result, providers have less time available to focus on the range of triggers and contributing factors for any given CRMC. This trend is unfortunate, as for many CRMCs the number and complexity of environmental and lifestyle triggers can be quite robust. For example, sleep changes have been described in ~50% of patients with migraine headaches, although 75% of patients also chose to sleep due to the migraine headache^5^. In addition, a study of 1207 patients with migraine headache identified no less than 16 possible triggers present in at least 5% of migraine sufferers^6^. A similar scale in triggers has also been noted for depression^7^, anxiety^8^, and chronic low back pain flares^9^. As such, tailored management of patients with these and other CRMCs often requires the provider take a detailed, longitudinal history with attention to temporal relationships—an approach that fits poorly with the practical constraints of modern clinical practice.

One approach that clinicians and innovators have advocated has been the use of technology to assist patients in self-management of CRMCs. In theory, through use of a mobile (mHealth) or web-based application, a patient could potentially self-manage his or her condition, incorporating both features specific to his/her pattern of CRMC recurrence, as well as general guidance from the treating provider. In light of this potential, mHealth applications have increased significantly in frequency over the years, with iOS apps including health and fitness groups increasing from 43k in 2013 to 98k in 2015^10^. Unfortunately, many of these approaches have failed to reach any meaningful level of adoption across the medical community^10, 11^, with most lacking any clinical validation prior to marketing to the general public.

In 2017, we formed the Individualized Data Analysis Organization (IDAO, analyzemydata.org) on the University of Colorado Anschutz Medical campus to guide a process for automation of self-management of CRMCs. The result was creation of a prototype iOS mHealth application called the iMTracker, which includes both a platform for patient-entered data to log recurrences of a given CRMC, as well as the opportunity to log possible triggers or suppressors of the CRMC, with an automated analysis providing both a data summary of the pattern as well as potential correlations for intervention (See Methods, below, for details). Through iterative use, patients can thus apply the iMTracker to identify and test lifestyle interventions (e.g., avoiding caffeine) toward the overall goal of reducing recurrence of their specific CRMC. The iMTracker is currently available in the iTunes store^12^ (Fig. 1), although clinical validation is required before use in medical settings.

**Figure 1.**
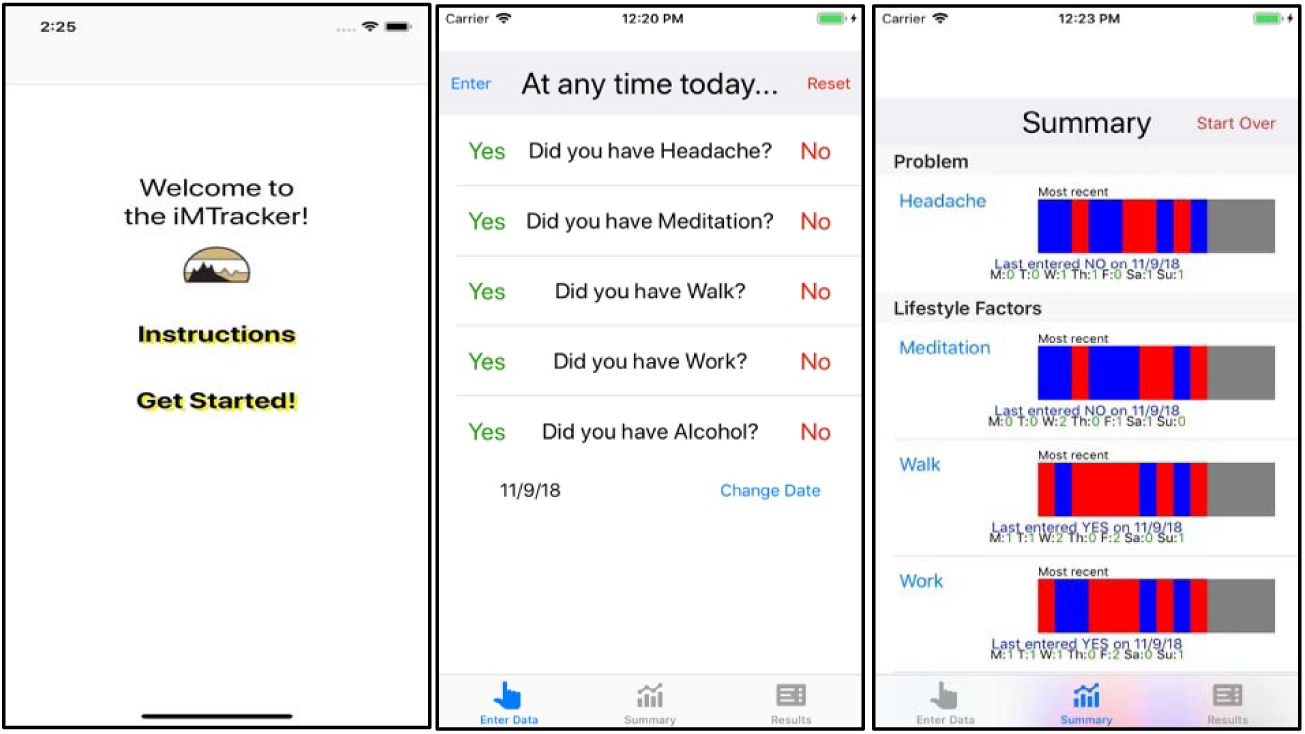
Screen shots of the iMTracker (iPhone)

In this investigation, we examined the potential applications and barriers to use of this iOS mHealth application in self-management of CRMCs.

## Methods

### Subjects

From May 15, 2019 until March 27, 2020, we recruited 153 subjects to pilot test the iMTracker app for iPhone for self-management of their chronic recurrent medical conditions using an internet-based study design. Inclusion criteria were age 18 or older, presence of a CRMC, and use of an iPhone. There were no official exclusion criteria, although based on study design and app functionality, subjects generally needed to be English-speaking and familiar with use of iPhone applications, as well as use of email and internet capability. We started with advertising using social media, such as Twitter, campus-based fliers, and provider word-of-mouth, but found limited recruitment, in which only two subjects were recruited. We then employed the TrialFacts patient-recruitment company (trialfacts.com, San Diego, CA, USA) to assist with internet-based recruitment. Subjects were provided a small financial stipend for participation. Written informed consent was obtained for all subjects. The study protocol was approved by the Colorado Multiple Institutional Review Board (CoMIRB, protocol #18–1000).

### mHealth App (iMTracker)

The platform of the iMTracker was designed based on an automated N-of-1 trial approach (Fig. 2) that includes both hypothesis generation and hypothesis testing, which can be built into the logic of an mHealth application. The iMTracker allows the user to select any problem (outcome) and any potential lifestyle factors (risk factors) or intervention that the user would like to test for an association with the outcome. Through iteration between hypothesis generation (i.e., ‘is there an association between risk factor A and occurrence of my condition?’) and hypothesis testing (i.e., ‘does changing risk factor A improve the rate of occurrence of my condition?’), the user is able to self-manage his or her condition towards an overall goal of reducing recurrence. Importantly, the overarching design of the iMTracker has been focused on application of edge computing^13, 14^ strategies that run on the mobile device itself, in order to allow complete usage of the iMTracker without need for transfer or storage of data on a server, which provides patients with a level of privacy and data security^15, 16^.

**Figure 2.**
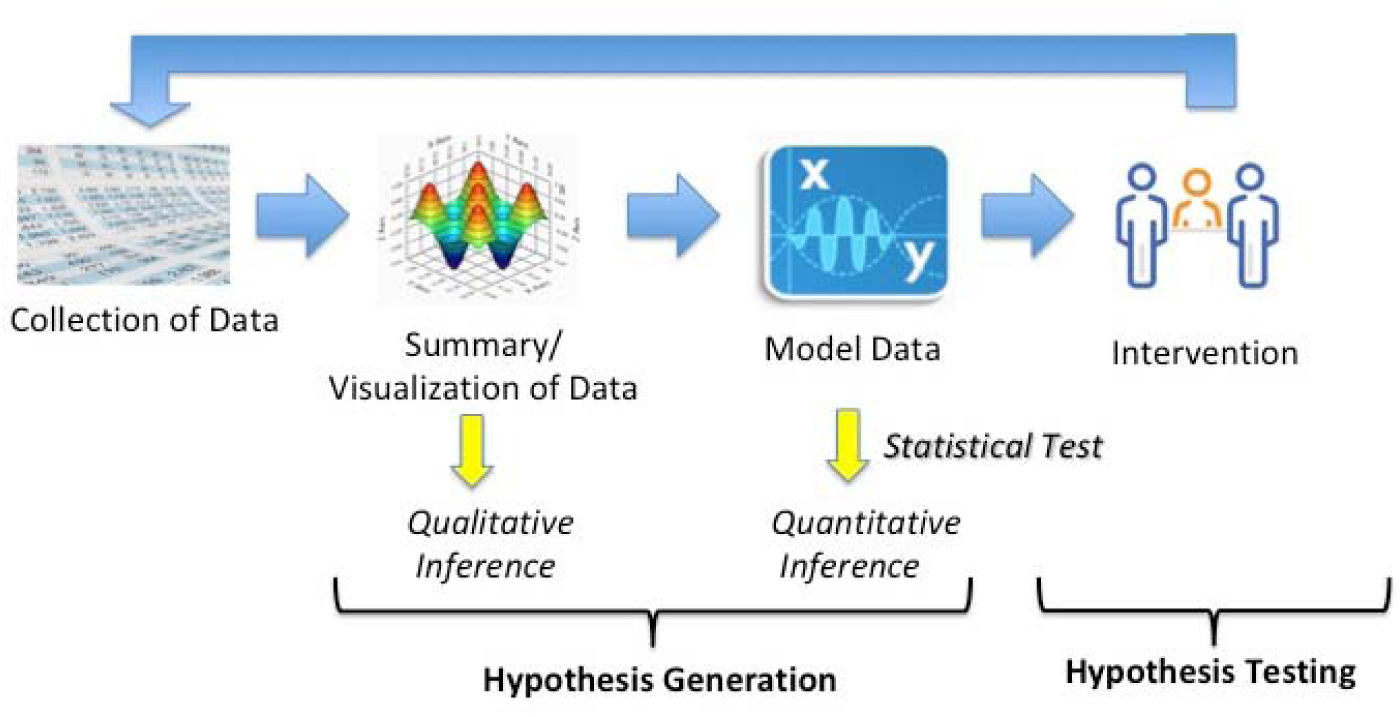
The automated N-of-1 approach motivating the iMTracker design.

Data is manually entered by the user, presented in a visual format, and then modeled for correlations between the selected outcome (problem) and potential risk factors (Fig. 1). Through built-in notifications, the iMTracker prompts the user to input data daily, and keeps a running summary of the inputs. The present analysis includes correlation between the outcome and risk factors on a daily basis and with one-day lag to identify risk factors that could potentially cause the outcome on the following day, using the phi statistic for correlation between discrete variables^17^. Although analysis is performed after only three days of data collection, users are informed that the accuracy of the correlation is higher with greater amounts of data collection. Once enough data has been collected to form hypotheses about causative associations, users are directed to reset the data collection and select an intervention in the form of a lifestyle modification, from which future data will examine the role of that intervention in reducing recurrences of the outcome.

Our goal in this study was to understand patterns and characteristics of the possible CRMCs and users themselves, as well as to identify obvious design and functional barriers to use of the iMTracker prior to use, prior to testing use of the device over a planned 3-month period. Future investigations will examine use patterns and additional design and functional barriers of the iMTracker.

### Survey

Our team designed a brief survey instrument with several goals in mind. First, we wanted to identify which specific CRMCs users selected for self-management using the iMTracker. These diagnoses were self-provided, and we did not perform separate validation with either treating clinicians or chart review. Second, we sought to collect information about the typical pattern of CRMCs—i.e., frequency of recurrence—in order to understand the burden of disease of a possible user of the iMTracker, and also to guide future work in automating analyses toward sufficient statistical power to detect associated lifestyle conditions and the effect of interventions. Third, we included questions aimed at detecting prior experience with regular data collection (e.g., ‘how often do you weigh yourself?’), information sharing (e.g., ‘how often do you post on social media?’), and electronic engagement with providers using email or secure messaging. Broadly, these questions helped to frame the users’ motivation for using this type of technology for self-management of CRMCs. Fourth, we inquired about concerns of using technology for self-management of conditions; specifically, we asked users to rank concerns related to data security, privacy, efficiency (time demand), and efficacy. Finally, we inquired about specific concerns with the iMTracker, and asked for qualitative input about design and function. In addition, we collected basic demographic information about categories of age, education, race, and ethnicity.

After informed consent was obtained, subjects were given a link to the online survey using a RedCap database. Subjects were guided through download and use of the iMTracker mHealth application for iPhone (iOS) by a member of the research team, and given the opportunity to provide qualitative feedback about app design, outside the survey data.

### Analysis

All study data was collected in a RedCap database. The analysis was performed using R, version 3.6.1 (R Studio, version 1.2.5019).

## Results

Surveys were completed by 135 subjects. Only two subjects were recruited by the study team outside of use of the TrialFacts company referrals. 131 subjects (97.0%) were under the age of 65 years old, with the predominant age range being 31–45 years old (65 subjects, 48.2%; Table 1). Most subjects had at least some college (98.5%), and most were Caucasian (80.7%) and non-Hispanic/Latino (90.4%).

**Table 1.** Demographics of iMTracker users.

| Age (years) |  | Education |  | Race |  | Ethnicity |  |
| --- | --- | --- | --- | --- | --- | --- | --- |
| Under 30 | 30 (22.2%) | Grade school only | 0 (0.0%) | Caucasian | 109 (80.7%) | Hispanic/Latino | 11 (8.2%) |
| 31 – 45 | 65 (48.2%) | High school diploma/GED | 2 (1.5%) | African-American | 7 (5.2%) | Not Hispanic/Latino | 122 (90.4%) |
| 46 – 55 | 27 (20.0%) | Some college | 35 (25.9%) | Asian | 7 (5.2%) | Unknown | 2 (1.5%) |
| 56 – 65 | 9 (6.7%) | College degree | 51 (37.8%) | American Indian or Alaskan Native | 5 (3.7%) |  |  |
| 66 – 75 | 4 (3.0%) | Master's degree | 40 (29.6%) | More than one/unknown | 7 (5.2%) |  |  |
| Over 75 | 0 (0.0%) | Doctorate degree | 6 (4.4%) |  |  |  |  |

The most common self-reported CRMCs that subjects planned to use the iMTracker to self-manage included pain (40.0%), which included low back pain and other musculoskeletal pain syndromes, followed by mental health conditions (17.8%), which included primarily anxiety and depression, and headaches (15.6%), Table 2. For most CRMCs, frequency was daily (40.0%) or multiple times a day (39.3%), with few occurring less often than monthly (3.7%), Table 2.

**Table 2.** User-selected CRMCs for application of the iMTracker. Note: GI = gastrointestinal conditions, including inflammatory bowel disease and irritable bowel syndrome.

| User-selected condition (CRMC) |  | Frequency of recurrence |  |  |  |  |
| --- | --- | --- | --- | --- | --- | --- |
|  |  | Multiple per day | Daily | Weekly | Monthly | Less than monthly |
| Pain | 54 (40.0%) | 29 (53.7%) | 21 (38.9%) | 2 (3.7%) | 1 (1.9%) | 1 (1.9%) |
| Mental Health | 24 (17.8%) | 8 (33.3%) | 12 (50.0%) | 3 (12.5%) | 0 (0.0%) | 0 (0.0%) |
| Headaches | 21 (15.6%) | 3 (14.3%) | 7 (33.3%) | 9 (42.9%) | 1 (4.8%) | 1 (4.8%) |
| GI symptoms | 7 (5.2%) | 0 (0.0%) | 4 (57.1%) | 3 (42.9%) | 0 (0.0%) | 0 (0.0%) |
| Hypertension | 7 (5.2%) | 3 (42.9%) | 2 (28.6%) | 1 (14.3%) | 0 (0.0%) | 1 (14.3%) |
| Palpitations | 5 (3.7%) | 1 (20.0%) | 3 (60.0%) | 0 (0.0%) | 1 (20.0%) | 0 (0.0%) |
| Dizziness | 2 (1.5%) | 2 (100%) | 0 (0.0%) | 0 (0.0%) | 0 (0.0%) | 0 (0.0%) |
| Other | 15 (11.1%) | 7 (46.7%) | 5 (33.3%) | 2 (13.3%) | 0 (0.0%) | 1 (6.7%) |
| **Total** | **135** | **53 (39.3%)** | **54 (40.0%)** | **20 (14.8%)** | **3 (2.2%)** | **5 (3.7%)** |

To assess willingness for daily data collection, we asked subjects how often they weigh themselves. 22 (16.3%) weighed themselves daily, 40 (29.6%) weekly, 34 (25.2%) monthly, 31 (23.0%) rarely, and 8 (5.9%) never. 47 subjects (35.3%) reported emailing or messaging with their primary provider regularly, with 51 (38.4%) reporting irregular/infrequent e-communication, 18 (13.5%) willing, but not needing e-communication, 9 (6.8%) not having the option, and 8 (6.0%) preferring in-person or over-the-phone communication. 39 (28.9%) of subjects posted on social media multiple times a day, 39 (28.9%) posted daily, 35 (25.9%) weekly, 10 (7.4%) monthly, and 12 (8.9%) posted rarely, if at all.

When asked about what factors were most concerning to subjects regarding use of an mHealth app to self-manage CRMCs, the most concerning was time needed to use the app (54.8%), followed by effectiveness/utility in self-management (43.7%), data security (25.2%), and data privacy (24.4%).

Finally, when asked how likely the subject was to use an mHealth app to self-manage their selected CRMC, 58 (43.0%) were very likely, 51 (37.8%) somewhat likely, 23 (17.0%) were neutral, 3 (2.2%) were somewhat unlikely, and 0 (0.0%) unlikely. Among the qualitative concerns raised by users specific for application of the iMTracker for self-management of their CMRC, design and functional limitations were generally noted, specifically related to the instructions and data entry process.

## Discussion

In this internet-based, pilot study of predominantly young and middle-age, educated, Caucasian subjects, we found that chronic pain, headaches, and mental health were the main CRMCs that subjects identified for self-management with an mHealth app. The frequency of recurrence of these conditions was high, with over 90% occurring at least weekly. Most subjects performed some degree of self-assessment/measurement as determined by up to monthly weight checks, most engaged in some form of e-communication with treating providers, and most posted regularly on social media. Not surprisingly, most subjects believed that an mHealth app could improve self-management of their CRMC; interestingly, the main concerns with an mHealth app was effectiveness and the amount of time needed to use the app, with less being concerned about privacy or data security.

The concern of subjects with effectiveness with overall app function is consistent with other studies of mHealth applications for self-management of CRMCs. In one investigation, it was found that only 3.4% of apps on the iTunes and Google Play stores promoted for management of depression and anxiety had research to justify their claims of effectiveness, with only 30.4% having expert input in development^18^. A study by Devan et al. of 19 apps available commercially for self-management of pain found only 2 that had been validated to improve health outcomes^19^. A similar lack of scientific support for commercially available mental health-targeted^20–22^ and pain^23, 24^ apps has been reported by other investigators. Although we did not inquire about prior use of mHealth apps for self-management, one can infer that most participants in this study had tried prior apps without success. Clinical validation of any mHealth app should be requisite before integration into the clinical care process, and our study further suggests that while users are optimistic that self-management using an app is possible, follow-up clinical studies will be needed.

Among the characteristics of the specific CRMCs that subjects identified for use of an mHealth app, recurrent pain, headaches, and mental health were highly represented. While these diagnoses were self-identified by users, and not validated with clinicians or clinical data (i.e., chart review), it does help to identify potential clinics and providers for testing mHealth apps. In addition, the majority of subjects noted a high frequency of recurrence of their condition, which is key in determining the number of subjects needed for a prospective study to demonstrate efficacy.

Although our study did not specifically examine adherence or efficacy of the iMTracker, the qualitative feedback provided early insight into the role of design and user-based feedback to guide improvements. Neuhauser et al., have previously noted that participatory methods linked with traditional health communication theory and methods can create effective health communication using artificial intelligence^25, 26^, highlighting the role of design science theory in development and refinement of mHealth applications. Such insights highlight the challenge that is unique to mHealth, and other health IT applications, in which consideration of user-based preferences and desires must be merged with information and guidance grounded in biology and evidence-based medicine principles. In terms of design life cycles, this requires an integrated design approach with features of both top-down (i.e., waterfall) strategies, as well as bottom-up, user-driven design (e.g., agile) strategies. Further work is needed to examine specific adherence rates of the iMTracker for self-management of CRMCs, although our team has already started to hypothesize how such processes can be integrated into the next generation of the iMTracker, including feedback from providers.

In addition to the data we collected in this investigation, equally important are the blind spots, or areas in which we failed to collect data. Specifically, the population we studied were primarily Caucasian, educated, and young/middle age adults; these were individuals who engage regularly with providers using technology, post to social media, and perform self-management with regular weight checks. Missing from our population are older subjects, subjects with less education, and subjects from underrepresented populations—the type of populations that have also been shown to have less close clinical follow-up for their conditions^27–29^, and whom might stand to benefit the most from an app that allows self-management. This population bias is critical in considerations of further app development as the design and functionality changes that would typically guide app development would be needed for successful integration with clinical care. Further work is needed on methods to include less represented populations in mHealth studies.

In conclusion, in this pilot study using internet-based recruitment, we found that potential early adopters of the iMTracker selected chronic pain, headaches, and mental health as preferred CRMCs for application, with relatively high (at least weekly) frequencies of recurrence. We also found clinical efficacy to be the primary concern with use of mHealth apps in general, with privacy and data security lesser concerns, and that efforts to examine and improve app efficacy and efficiency will be critical to successful integration with clinical care. We also identified population bias in the subjects enrolled using internet-based recruitment alone, and note that additional efforts will be needed to ensure that future studies enroll sufficient numbers of underrepresented populations, specifically older, non-Caucasian, and less education populations.

## Data Availability

Data will be made available upon request and approval by the University of Colorado Multiple IRB (COMIRB)

## Acknowledgments

Enrollment in this study was performed with assistance from TrialFacts (TrialFacts.com; San Diego, CA). This research was supported by grants from the NIH/NHBLI (MR: K23HL127296).

